# Pre-chronic kidney disease -- Serial creatinine tracks glomerular filtration rate decline above 60 mL/min

**DOI:** 10.1101/2024.09.17.24313678

**Authors:** Cyril O. Burke, Leanne M. Burke, Joshua Ray Tanzer

## Abstract

**OBJECTIVES:** Primary care for chronic kidney disease (CKD) stage 1-2 has recently been shown to slow CKD progression. We describe an approach to detect early decline in glomerular filtration rate (GFR) above 60 milliliters per minute (mL/min), before CKD stage 3.

**METHODS:** We re-examined a standard reference that found low tubular secretion of creatinine (TScr) at GFRs above 80 mL/min and suggested “…observation of subtle changes in serum creatinine levels”. We explain why that method extends down to 60 mL/min and summarize why estimated GFR (eGFR) is unreliable above 60 mL/min.

**RESULTS:** Four patient cases show how serum creatinine (sCr) referenced to an individual’s historical maximum suggests increased risk, triggering investigation to separate benign processes that alter sCr from decline in GFR of prechronic kidney disease (preCKD).

**CONCLUSIONS:** At GFRs above 60 mL/min, serial creatinine is more reliable than GFR estimating equations and appears practical for ‘race-free’ clinical monitoring and early intervention.

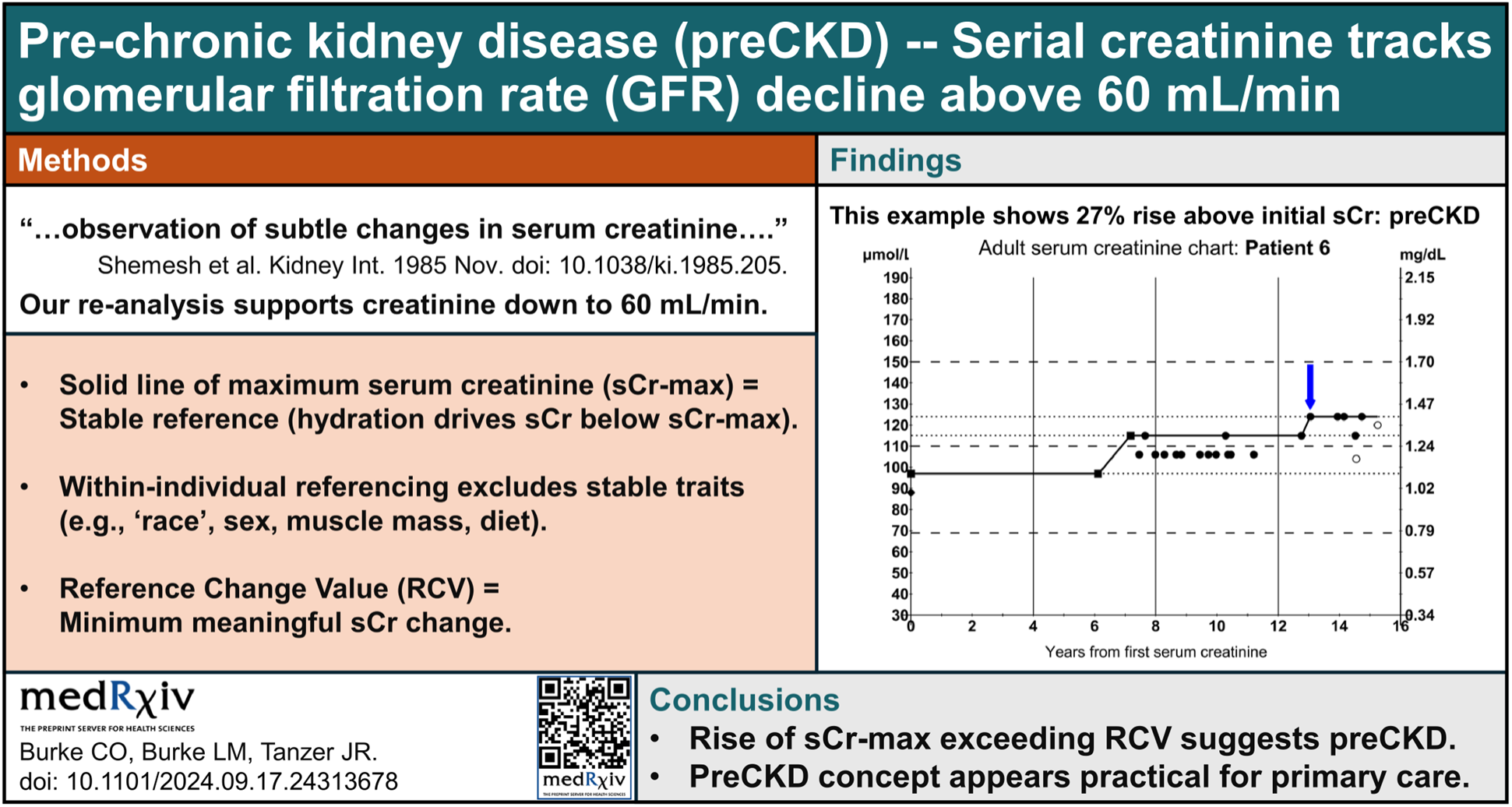

**STRENGTHS OF THIS STUDY:**

1. Includes serial creatinine proof-of-concept cases that show preCKD
2. Reviews ‘race-free’ properties that favor serial creatinine for early kidney disease
3. Addresses mathematical limitations of GFR estimating equations
4. Re-analyzes a 40-year-old standard reference that still influences research, showing its overlooked importance for CKD stage 1-2

## 1. INTRODUCTION

**Note:** An eminent European nephrologist wrote, “The thesis of the authors, that using the baseline serum creatinine of a given patient would potentially improve the earlier diagnosis of kidney disease, even in the normal range, is in line with the experience of this reviewer, who always retrieves, whatever the difficulty of reaching that goal, past results of blood tests, and uses them as a way to date the onset of kidney disease, sometimes with important prognostic implications.”

We expanded our article in response to reviewer suggestions (see four rounds of review in the comments) and eventually divided our research between this proof-of-concept study for early diagnosis of kidney disease and another, re-analyzing APOL1 kidney disease, that found support for new ethics of ‘race’ [1].

Between 1990 and 2023, global chronic kidney disease (CKD) rose from 27^th^ to 9th leading cause of death: “The global age-standardised prevalence of CKD in adults was 14·2%…. Most people had stage 1–3 CKD, with a combined prevalence of 13·9%” [2].

### PreCKD is primary care

Primary care research is essential for diagnosing early-stage CKD. Primary care clinicians manage GFRs down to 30 mL/min [3,4,5] because CKD exceeds the availability of nephrology specialists [6]. GFR suggests CKD when approximately halved to 60 mL/min (equivalent to functional loss of one kidney) and prompts nephrology referral when halved again to 30 mL/min (equivalent to functional loss of 1.5 kidneys).

Primary care measures to slow progression of CKD include drugs (e.g., ACE inhibitors, GLP-1s) and case managers to monitor kidney function and provide education [7]. However, most adults with CKD remain undiagnosed [8,9] because mathematical limitations of GFR estimating equations preclude diagnosis of CKD stage 1-2 (i.e., at GFRs above 60 mL/min) without another biomarker (e.g., proteinuria) [6].

The sCr ‘normal range’ (reference interval or population variation) is much broader than any person’s usual within-individual variation, so detecting early CKD without a non-creatinine biomarker (e.g., proteinuria) is challenging.

Shemesh et al recommended “…observation of subtle changes in serum creatinine levels” at GFRs above 80 mL/min (while also suggesting limitations of creatinine at GFRs below 80 mL/min and insensitivity of its population reference interval—often called the “creatinine-blind range”) [10]. Re-analysis of their data (see **Appendix: 5.2 A standard reference**) validated extension of that recommendation down to 60 mL/min.

We present a method to detect GFR decline at the top of the primary care range—above 60 mL/min—where sCr is within its reference interval and eGFR is unreliable. Tracking within-individual change of sCr has been shown to increase detection of acute kidney injury (AKI) [11,12,13] and correlate with CKD progression [14,15]. Charting sCr can also reveal ‘preCKD’, which is like prediabetes [16] and prehypertension [17], but because serum glucose and blood pressure have smaller between-individual variations, those conditions do not <u>re</u>q<u>uire</u> within-individual comparison.

## 2. RESULTS

### 2.1 Diagnosing preCKD

#### 2.1.1 Definitions

#### Maximum creatinine—sCrMax

Charting sCr levels over time (from first known sCr) creates a cluster of datapoints that reveals an individual’s “normal range”. While the cluster’s lower limit may vary with vigorous hydration, its upper limit—the maximum sCr (sCrMax)—approximates minimum GFR to that date (e.g., during fasting or dehydration). Its Y-axis span reflects combined analytic variability (within and between laboratories) and individual biologic variability due to muscularity, diet, and medicine effects.

When a new creatinine result exceeds the sCrMax, successive increments push the sCrMax higher until it is stable (for a time), making the accuracy and precision of assays and variability within and between laboratories less critical—sCrMax simply rises a little higher to encompass the variability in that patient. However, a subsequent significant increase in sCrMax possibly represents a true decline in GFR.

#### Reference change value—RCV

The sCr reference change value (sCrRCV) represents the minimum meaningful sCr change, which varies with physical activity: sCrRCV is 13.3% in healthy sedentary individuals and 26.8% if physically active [18]. A between-laboratory sCrRCV was reported [19], but because the sCrMax reference point equals the peak of all previous sCr values, a new sCr exceeding all prior values by >sCrRCV more likely represents a true decline in kidney function to preCKD.

#### PreCKD

In **Fig 1**, we divided guidelines [20] for GFRs between 120 and 60 mL/min into three subgroups: **(1)** stable ‘normal’ function (above 90 mL/min, not shown); **(2)** CKD stages G1 (>90 mL/min) and G2 (90-60 mL/min) with other identifying abnormalities (e.g., proteinuria); and **(3)** significant rise in sCrMax, which may be either reversible impairment (i.e., not CKD) or early CKD without another biomarker—preCKD.

**Fig 1.**
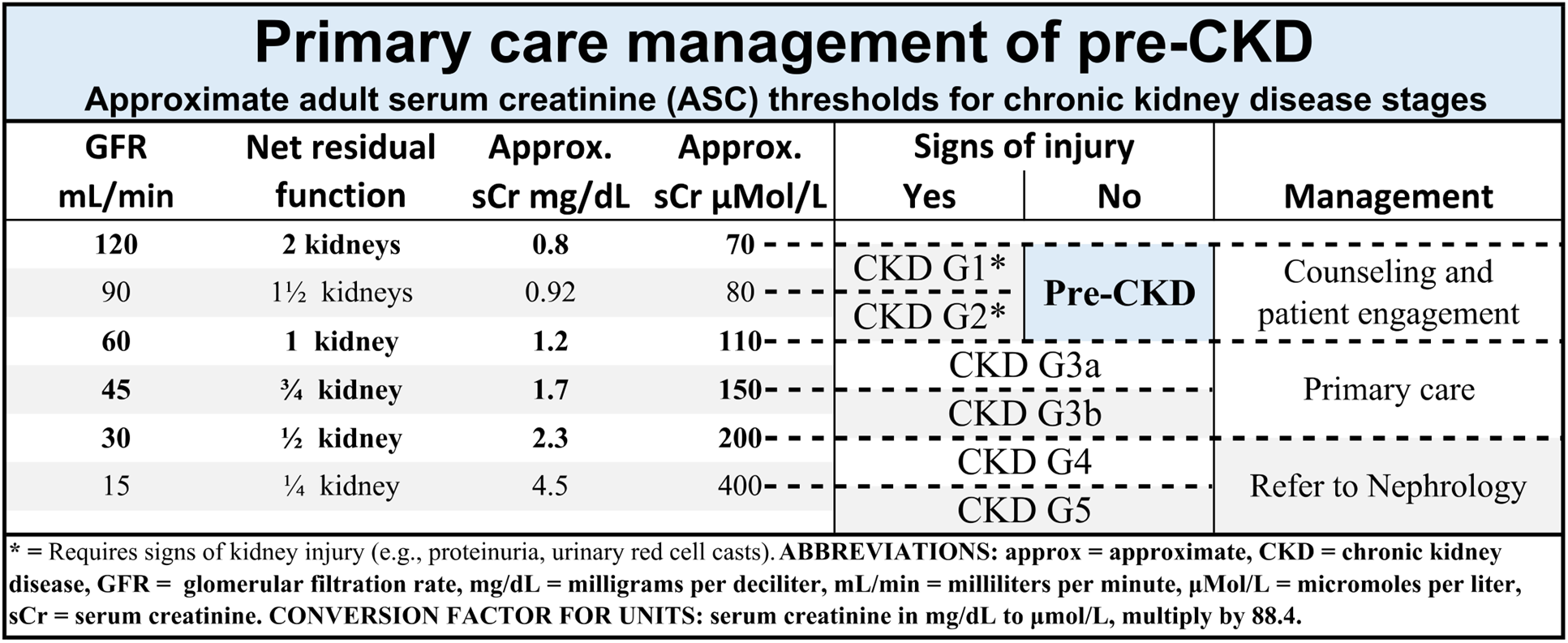
Staging early kidney impairment. The sCr boundary values are rough approximations (due to marked population variation between individuals) but show that seemingly small increases in sCr can signal significant decline in GFR and loss of kidney function.

**Fig 1** highlights the potential significance of small changes in sCr, with approximated sCr thresholds based on the fitted curve in **Appendix: 5.2.2** and rough error margins. However, these example thresholds lack specificity for individual care because they were derived from cross-sectional population data, with broad population variation especially above 60 mL/min (preCKD range). Observing within-individual sCrMax for change that exceeds sCrRCV offers individually actionable sCr limits for clinical use.

### 2.2 Selection of cases

From August 2019 to November 2021, in a non-nephrology specialty practice, we prospectively identified and collected historical laboratory data on patients with sCr ≥1.0 mg/dL and without diagnosed kidney disease. We included a “normal” patient (Patient N) for comparison. All had primary care providers and Medicare or private health insurance. Consulted nephrologists were unaware of the pattern of historical serial sCr levels collected at the time, which were analyzed later when defining the adult serum creatinine (ASC) charts. Patients who either were referred back to their primary clinicians or declined ongoing care were ‘lost to follow up’, and were not contacted thereafter.

The 11 patients listed in **Fig 2** included seven White, three Black, and one Asian American-Native Hawaiian/Pacific Islander. The median age was 73 for White and 65 for Black patients. Hypertension was present in 10 of the 11 patients. Diabetes or pre-diabetes was noted only in the three Black patients.

**Fig 2.**
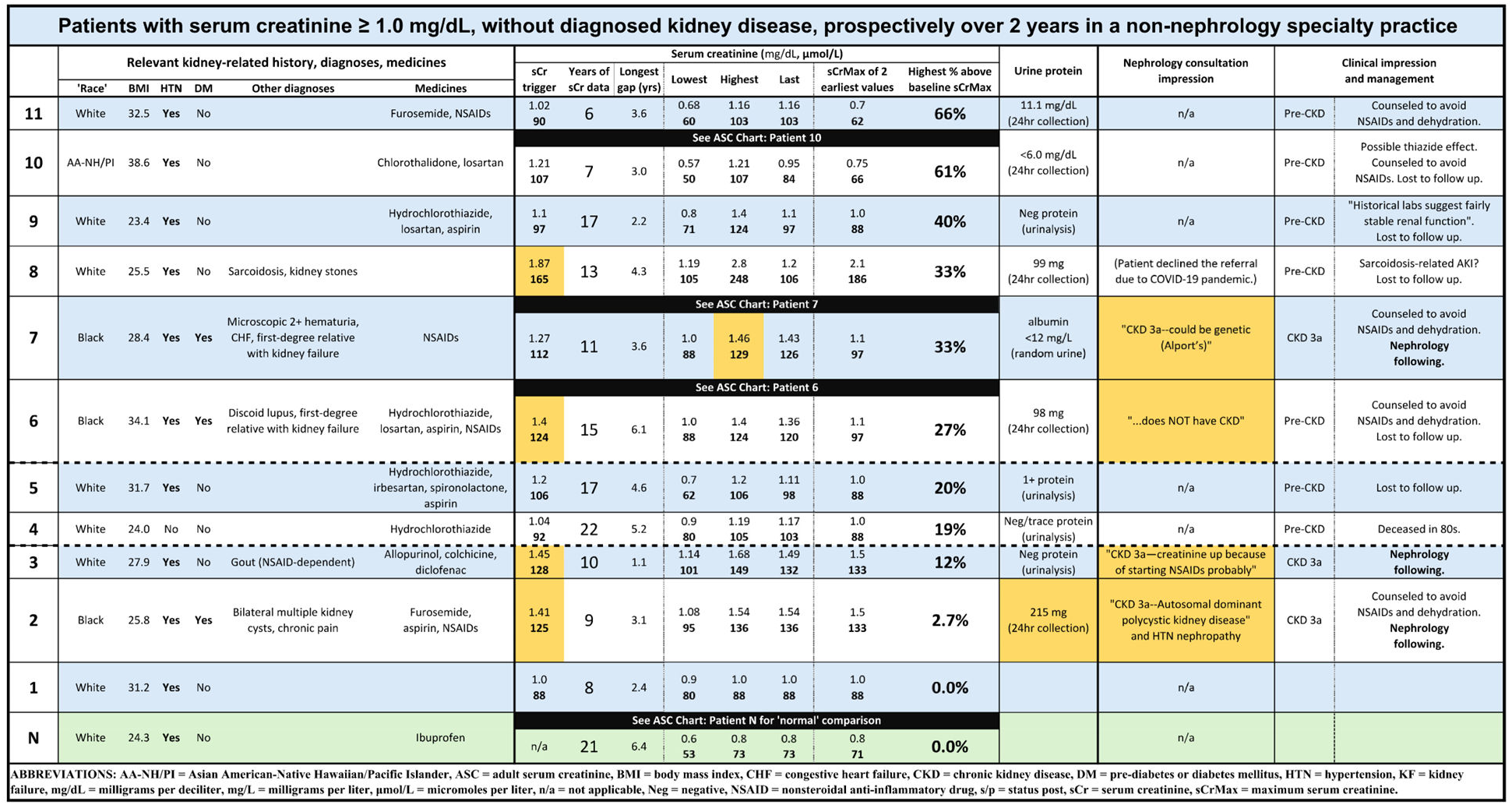
Patients with serum creatinine ≥1.0 mg/dL, no kidney diagnosis, in a non-nephrology specialty practice. Highlighted (orange) cells indicate results that prompted referral to nephrology and impressions of the various nephrologists. Dashed lines represent boundaries of serum creatinine reference change value (sCrRCV) for “change relative to sCrMax”: 13.3% (lower) in healthy sedentary individuals and 26.8% (upper) if physically active patients. For comparison, Patient N, in green, had stable medical issues and kidney function over many years.

Median availability of sCr data was 11.6 years (mean 12.8, range 4.4 to 21.0 years), see supporting **S1_dataset**. The higher of two earliest sCr values served as baseline sCrMax for calculating future sCrMax and changes in sCr. In **Fig 2**, four White patients (mean age 72.3) had sCr too low for nephrology referral. **Patient 1** had peak sCr 1.0 mg/dL and relative change 0.0%. **Patients 4, 5, and 11** met preCKD criteria, with relative change from 19% to 66%, but highest sCr only 1.16 to 1.2 mg/dL.

Six patients had sCr ≥1.4 mg/dL. Of three White patients (mean age 75.3), two were referred to nephrology: **Patient 3** had gout dependent on nonsteroidal anti-inflammatory drugs (NSAIDs); **Patient 8** had sarcoidosis but declined nephrology evaluation (sCr range: 1.19-2.8 mg/dL). The third, **Patient 9**, was octogenarian, with “fairly stable sCr” that regressed (sCr range: 0.8-1.4 mg/dL).

Three Black patients (mean age 66.7) were referred to nephrology: **Patient 2** was diagnosed with “polycystic kidney disease” (sCr range: 1.08-1.54 mg/dL, relative change 2.7%—urine protein 215 mg/24hr, renal cysts by computed tomography); **Patients 6 and 7** are described below, in **Section 2.3**.

Even though all the patients in **Fig 2** showed a gap of several years in sCr measurements, early diagnosis of kidney disease may be possible even from limited longitudinal sCr data. In a non-nephrology specialty practice, the small number of cases identified over two years suggests the specificity of sCrMax, sCrRCV, and serial sCr for early kidney disease may be a practical method for primary care clinicians to identify a subset at higher risk.

### 2.3 Longitudinal creatinine

As examples, we present charts for three of the 11 patients with sCr ≥1.0 mg/dL and one “normal” patient, with stable sCr <1.0 mg/dL. The ASC charts show four dashed horizontal guidelines: the lowest at 68.9 μmol/L (best sensitivity and specificity for increased CKD risk in men [21]) and three spaced at 40 μmol/L intervals—not meant as absolute boundaries (due to wide population variation) but approximating lost kidney function from small changes in sCr.

#### 2.3.1 Stable for decades

**Patient N** (“normal”) was White, on no relevant medicines, with stable diet, muscle mass, physical activity, and normal-range sCrMax for 21 years, **Fig 3**. Plotting sCr over time provided more reliable confirmation than isolated sCr levels.

**Fig 3.**
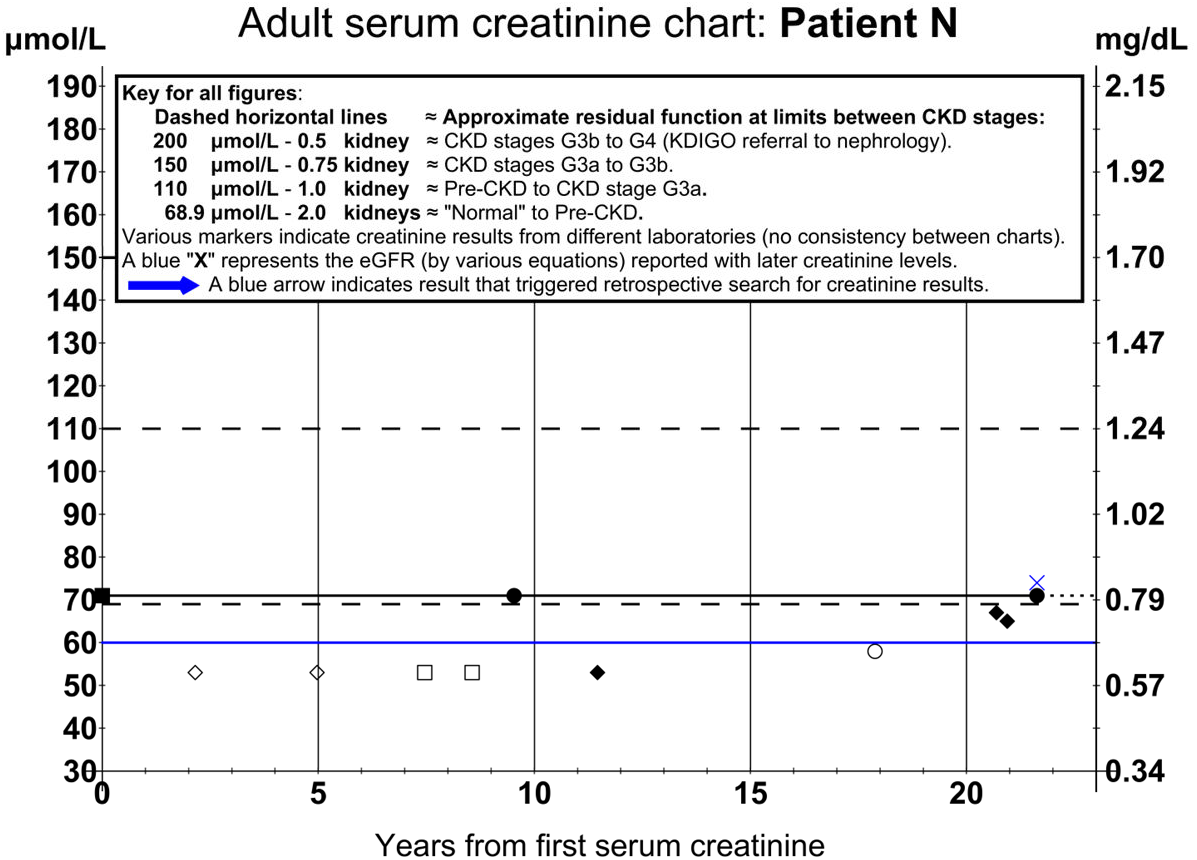
Stable for two decades. Patient N (“normal”). This adult serum creatinine (ASC) chart shows a six-year gap in sCr testing but stable maximum sCr (sCrMax) for 21 years. **Note:** The sCr levels below the line of sCrMax represent improved glomerular filtration (e.g., after hydration), varying within a range that may reflect renovascular adaptive capacity. The sCrMax line smooths the limit of apparent minimum glomerular filtration capacity.

#### 2.3.2 High-risk, stepwise progression

**Patient 6** was Black, ‘muscular’, with stable diet and activity for 15 years. Serum creatinine of 1.4 mg/dL prompted nephrology consultation. Back-referral concluded, “muscular build…. elevated serum creatinine level—normal renal function…. does NOT have CKD”.

Relative to sCr at time zero, increases at years 7 and 13 totaled 27.8%, **Fig 4**, exceeding ‘physically active’ sCrRCV. Urine albumin-creatinine ratio, 16.67 μg/mg Cr, and 24-hour urine protein-creatinine ratio, 98 mg/g Cr, appeared normal, but high urine creatinine (1,905 mg/24 hours) can mask non-nephrotic proteinuria [22,23], suggesting possible CKD stage A2 [24]. The preCKD pattern prompted counseling and referral to endocrinology to optimize diabetes care.

**Fig 4.**
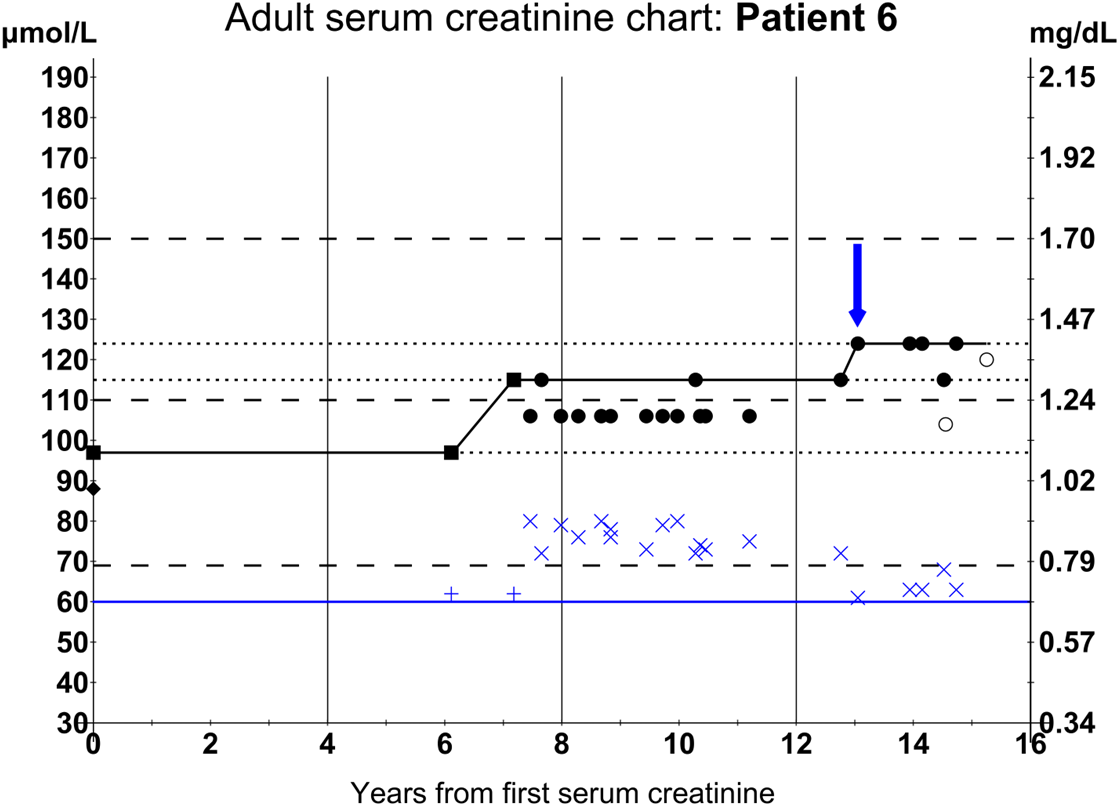
High pre-test probability, stepwise progression. Patient 6. The ASC chart shows a six-year gap in sCr testing and stepwise progression in sCrMax, suggesting two periods of injury, at years 7 and 13. See Fig 3 for an explanation of ASC chart construction.

#### 2.3.3 Familial and environmental

**Patient 7** was Black. Relevant medicines included NSAIDs. Nephrology consultation suspected genetic nephropathy (Alport syndrome).

The sCr chart showed progressive rise in sCr of 33%, **Fig 5**, possibly worsened by NSAID use. Horizontal lines indicate stable sCrMax from 0 to 3 years and 7 to 8.5 years. Sloped lines indicate increasing sCrMax (i.e., without intervening measurements), sometimes over prolonged intervals, suggesting the need for guidelines on minimum frequency of sCr sampling.

**Fig 5.**
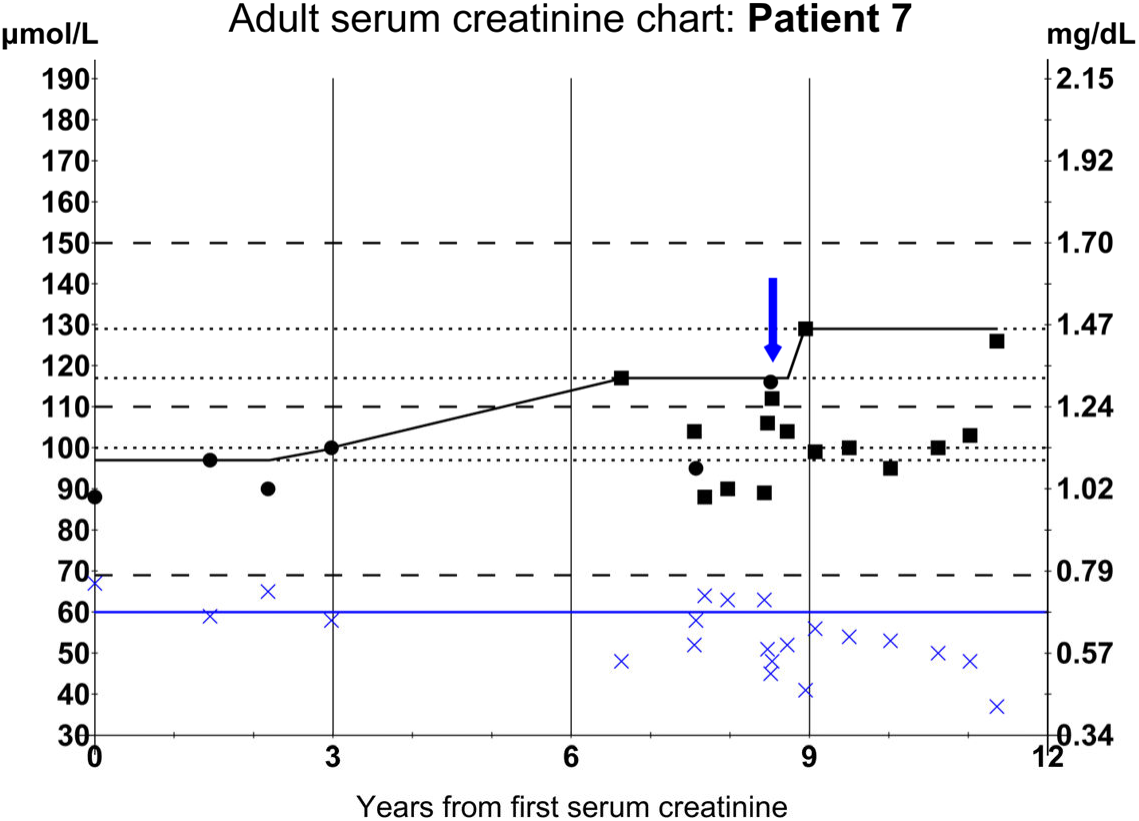
Familial and environmental factors. Patient 7. The ASC chart shows a three-year gap in sCr testing and progressive increase in sCrMax. See Fig 3 for an explanation of ASC chart construction.

#### 2.3.4 Thiazide holiday

**Patient 10** was Asian American-Native Hawaiian/Pacific Islander, with hypertension. The inciting sCr level (blue arrow) suggested significant loss of kidney function. The 24-hour creatinine clearance (CrCl) was 84 mL/min, without proteinuria.

The sCrMax rose progressively over 11 years (solid line), **Fig 6**. The drops in sCr during two intervals when thiazide diuretic was stopped for hyponatremia (then resumed for ankle edema) or reduced (arrow) suggested thiazide altered kidney function and potential for trial off thiazides before invoking preCKD or nephrology consultation.

**Fig 6.**
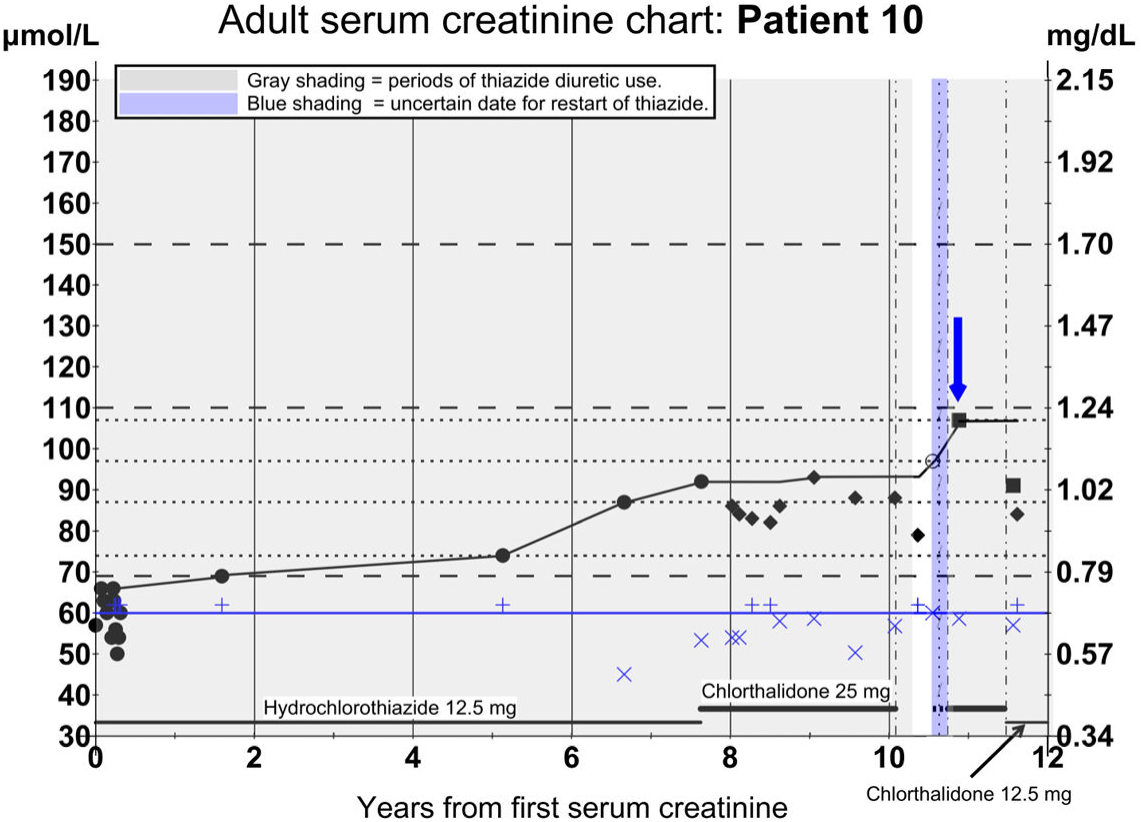
Chlorthalidone holiday. Patient 10. The ASC chart shows a four-year gap in sCr testing and a progressive increase of sCrMax over eleven years. Gray zones represent treatment with a thiazide diuretic. The five-month clear zone represents sCr without thiazide bias—off due to hyponatremia. The blue zone indicates imprecise date of resuming thiazide diuretic for ankle swelling. See Fig 3 for an explanation of ASC chart construction.

## 3. DISCUSSION

### 3.1 Early diagnosis

The patient cases show early diagnosis using within-individual referencing of longitudinal sCr.

### 3.2 Why sCr works in preCKD

#### 3.2.1 Optimal clinical use

Lab tests should provide optimal clinical use, regardless of agreement with highly variable physiologic parameters, like measured GFR (mGFR). For preCKD, sCr is a convenient, stable, and almost ideal endogenous filtration marker that explained virtually all eGFR variability in longitudinal studies [25]. Within-individual change gives superior decision-making for preCKD, versus a mGFR or a newly modified eGFR, eliminating the need to tolerate an enormous (±30%) “error”. Of two physiologic limitations to monitoring GFR with creatinine [26], sCr is affected by one: tubular secretion of creatinine (TScr), which is acceptably low at preCKD GFRs (see **Appendix: 5.2 A standard reference** for detailed justification).

#### 3.2.2 PreCKD urine flow

Urine collection is simpler at preCKD urine flows. Without the complexities of exogenous filtration markers (see **Section 3.3**), ‘quick CrCl’ may have acceptable performance under some conditions [27]. Above 60 mL/min, GFR changes can signal marked increase of “augmented renal clearance” [28,29,30], which could mask a decline into preCKD.

#### 3.2.3 Low Index of Individuality

Despite larger population variation (between individuals), sCr has both low analytical variation and low within-individual variation [31,32]. Its low Index of Individuality favors comparison to an individual’s baseline sCr rather than a population reference range [33,34], which should also be true for values derived from sCr, including eGFR. Creatinine’s low Index of Individuality undermines one goal of eGFR equations—to offer universal population reference intervals, which remains elusive despite decades of incremental adjustments.

The problem with population reference intervals is less noticeable after entering CKD stage 3, both because the effects diminish as GFR falls with advancing CKD and these smaller shifts mainly occur under the expert care of nephrologists. However, where early CKD overlaps normal kidney function, false-negative results have major consequences by masking awareness of possible kidney disease.

False-positive results also have consequences. A decision analysis by den Hartog et al concluded that the benefit of eGFR over sCr was reversed with even minimal reduction in quality of life from incorrect diagnosis due to markedly more false-positive diagnoses of CKD with eGFR than with sCr alone [35].

#### 3.2.4 Makes ‘race’ and other confounders irrelevant

Within-individual sCr reference makes ‘race’ irrelevant, mitigating one source of disparities in kidney care [36] and setting research expectations for similar rates of CKD and KF, regardless of ‘race’ [1].

Within-individual sCr reference also minimizes effects of age, sex, and (if stability can be confirmed) individual traits (see **Appendix**: **5.3 Classic creatinine cofactors**). Primary clinicians must exclude non-renal influences on sCr, including acute illness; changes in activity [37], BMI, meat ingestion [38,39], fasting, or dehydration [40]; and drug effects from NSAIDs, thiazides, and even nephroprotective treatment using ACEI/ARB and SGLT2-I classes. In athletes, the sCr effect varied by sport and was transient, suggesting that periods of training, competition, and recovery should be assessed separately [41].

Although touted as superior to sCr, serum cystatin C (sCysC) is affected by inflammation, corticosteroid use, diabetes, age, height, weight, smoking status, concentration of C-reactive protein, and cystatin C gene variants [42,43].

### 3.3 eGFR drawbacks above 60 mL/min

Comparing CrCl to the mGFRs shows why sCr is potentially useful. Because mGFR has population variation as wide as sCr, with much greater individual physiologic variability compared to the relatively stable sCr and sCysC [31,44], eGFR equations have wide error margins that often overlap the CKD stages. The eGFR equations (Modification of Diet in Renal Disease, CKD Epidemiology Collaboration, others) cannot eliminate this overlap. Consequently, eGFR has poor predictive value at GFRs above 60 mL/min (often misinterpreted as “absence of kidney disease”).

At sCr below 1.0 mg/dL (GFR above 60 mL/min), the inverse of sCr in the calculation amplifies differences between the calculated eGFR and the mGFR, contributing to wide “P30” error margins (i.e., percentage of estimates within ±30% of the reference value). A currently acceptable P30 of 85% means that an eGFR of 60 could represent an mGFR between 42 and 78 mL/min, with 15% of eGFRs falling outside even that broad range.

Porrini et al examined eGFR equations from the 1950s to the present and identified the mismatch between eGFR and mGFR: <u>regardless of the analyte</u> (sCr, sCysC, Beta Trace Protein), eGFR classifies the stage of CKD differently in 30 to 60 percent of patients [45], especially for preCKD. Similar misclassification was shown for potential kidney donors [46], pre-dialysis care [47], and recent “race-free” eGFRs [48,49].

Farrance et al showed uncertainty in eGFR is mathematically unavoidable [50]—neither a switch to sCysC nor a panel of tests [51] is likely to remedy them. Kallner noted, “the larger uncertainty associated with the eGFR will emphasize the sensitivity of S-Creatinine in relation to eGFR” [52].

Although within-individual referencing of eGFRs could suggest early decline in GFR, accumulated inaccuracies in eGFR make it especially misleading for preCKD, even when a baseline is available. Compared to sCr alone, in the preCKD range, there appears no need to introduce the mathematical errors and uncertainties of converting sCr to eGFR. Direct, within-individual comparison of sCr by direct analytical measurement is mathematically cleaner, and preferable for preCKD, versus manipulating calculated eGFR “quantity values” that accumulate uncertainties of the equations and their inputs [50].

#### 3.3.1 True GFR

True GFR is a dynamic variable, constantly changing with variation in physiologic factors like blood pressure, sympathetic autonomic activity, and volume status (e.g., volume expansion from intravenous infusion or contraction from “dehydration”).

Because compensatory mechanisms blunt the decline in GFR by increasing pressure, perfusion, and filtration in remaining nephrons, even ‘true’ GFR is not strictly proportional to perhaps the ultimate measure of CKD: loss of functional nephrons, which cationic ferritin-enhanced MRI (CFE-MRI) might eventually reveal [53,54].

#### 3.3.2 Gold standard vs reference standard

Because GFR cannot be measured instantaneously or directly, it has no <u>true</u>, 100% sensitive, 100% specific “gold standard” (GS). Lack of a GS precludes having an imperfect GFR reference standard with <u>known</u> diagnostic accuracy. While some regard inulin clearance as a GS, we do not agree for this reason: like CrCl, <u>all</u> the mGFRs are simply clearances of a filtration marker, with the exogenous mGFRs subject to significant additional complexities [26,81,55]. The common assertion that injecting inulin or another filtration marker for mGFR is the GS overlooks unverifiable assumptions in their various methods [56], which cannot be validated. There are also physiologic, within-individual variations that potentially alter GFR (e.g., by expanding volume). Thus, exogenous mGFRs are mutually inconsistent [57,58,59,60] and have misclassification errors [61].

Limiting observations to preCKD range, creatinine appears at least comparable to inulin and superior to other filtration markers, **Fig 7**.

**Fig 7.**
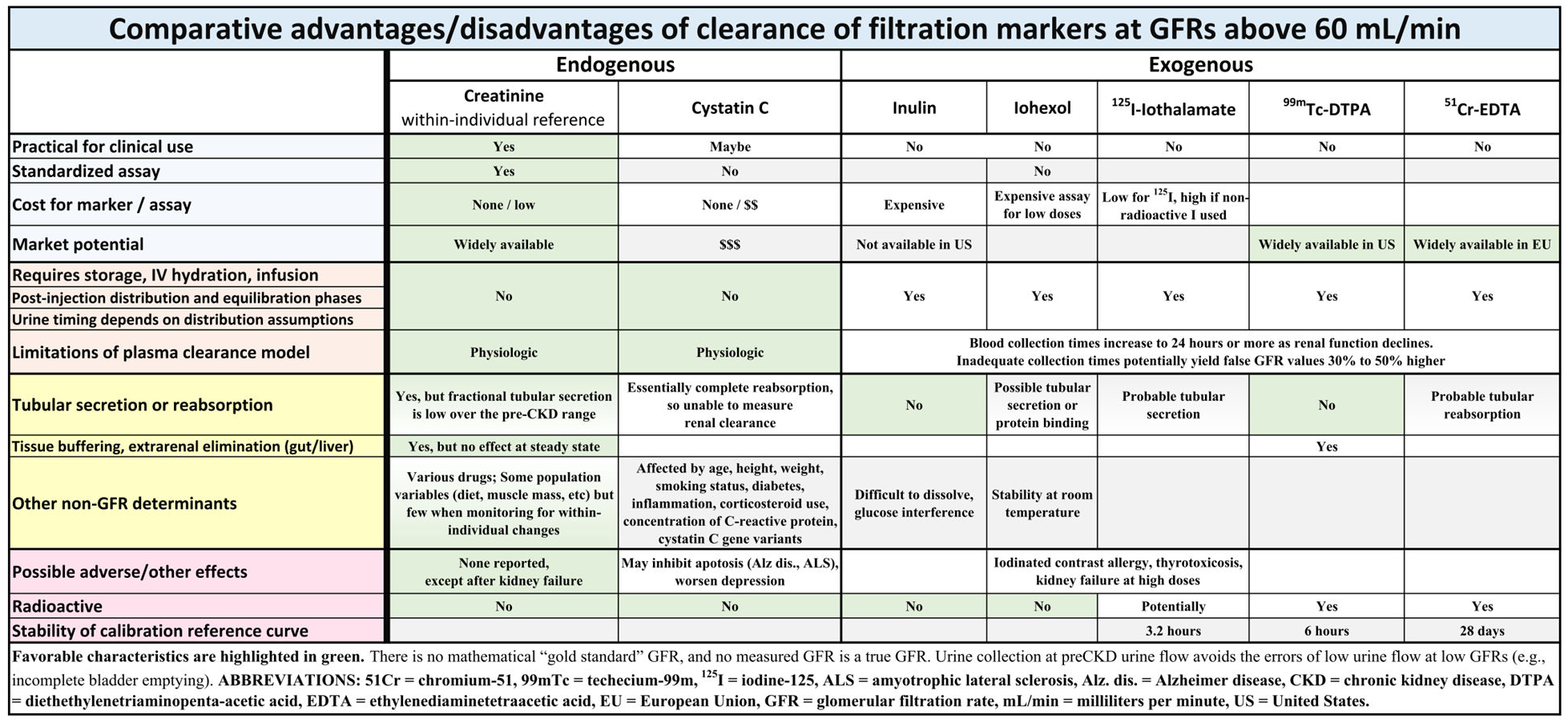
Comparative advantages/disadvantages of filtration markers, especially at GFRs above 60 mL/min. Favorable characteristics are highlighted in green.

Bland-Altman plots of two “field-methods” properly put their <u>difference</u> on the Y-axis against their <u>mean</u> on the X-axis when both are sources of error [62,63]. Plotting a <u>true</u> GS on the Bland-Altman X-axis makes sense [64], but studies often plot mGFR on the X-axis [44], wrongly attributing all error to creatinine [65,66,56,10]. Bland-Altman plots demonstrate challenges in properly interpreting numeric information that has limits on the values it can assume. Cross sectional data are not equipped to represent this dynamic system. We noted influences of urine volume, sCr, and value of the denominator, which could explain these patterns of relationships. Even today, collecting data on time-specific dynamic systems like this would be challenging. We offer this as an explanation of how limited methods could result in systematic problems with interpreting lab tests, but more research is needed to understand these relationships in more detail.

#### 3.3.3 Validity and reliability

The validity of clinical measurements is context-dependent, so more important than identifying an unobtainable GS, the focus should be on the context in which obtainable measurements are valid indicators of GFR in CKD patients.

For this reason, we emphasize the importance of examining sCr over time. Even if another mGFR had better reliability and concurrent validity as an estimate of GFR at a single point of measurement, CKD by definition is longitudinal, so a limitation of interpreting a single measurement would be overcome by interpreting a pattern of results within the same individual.

Furthermore, given the availability of sCr relative to other mGFRs, sCr is a more practical indicator of CKD condition so long as the interpretation is informed by any identified limitations in reliability. As our cases show, a valid inference can still be drawn from a p<u>attern</u> of increasing sCr values within the same individual over time. This pattern may already be evident in historical sCr results, which could be stored in personal devices for access during acute care.

With fractional TScr 16% over the preCKD range, screening with creatinine is an imperfect reference standard with known diagnostic accuracy. This suggests that clinical validation may be most appropriate to assess the predictive value of longitudinal changes in sCr [67]. It also suggests that interpretive parameters will be clarified with experience gained over time by validation studies [68], which may appeal to primary care clinicians.

Finally, in addition to clinical validity, Kassirer advocated weighing clinical utility—expense, risks, invasiveness, availability, and practicality for clinical use [69]. At preCKD GFRs, all favor creatinine.

## 4. CONCLUSION

At GFRs above 60 mL/min, direct measurement of serum creatinine (sCr) offers a ‘race-free’ approach to detect early kidney impairment that avoids the limitations of eGFR quantity values.

After excluding benign causes (like diet, medicine effect, muscle mass), primary clinicians interpretating longitudinal changes in sCrMax may achieve early diagnosis of preCKD.

In a non-nephrology specialty practice, an incidental sCr 1.0 mg/dL or higher triggered limited investigations for preCKD. Further research may show an optimal change in sCr for clinical use and whether monitoring changes from a youthful sCrMax will allow earlier preventive measures that encourage patients to avoid kidney risks.

The relatively low cost of sCr assays raises hope for increased availability [70] and international comparisons under different environmental and social conditions to reveal preCKD cofactors, improve health, save resources, and reduce CKD disparities.

## Data Availability

All data produced in the present work are contained in the manuscript and supporting files.

## 5. APPENDIX

## 5.1 Ethics statement

### ‘Race’ in research

We do not present ‘race’ as an input to clinical decisions, a means of stratifying care, nor to hypothesize genetic difference without data [71].

### Human subjects

The data presented in patient cases were obtained for non-research purposes in a direct-treatment relationship that was not considered human subject research. Brown University Health (formerly Lifespan Health System) Human Reseach Protection Program waived ethical approval for this work.

### Privacy

Our de-identified cases are not protected health information and required neither patient consent nor review or waiver by a research ethics board [72].

## 5.2 A standard reference

Shemesh et al used cross-sectional sampling of 171 patients (i.e., population variation, rather than individual variation) to dichotomize a continuous physiologic process and suggest limitations of creatinine as a filtration marker and insensitivity of its population reference interval (often called the “creatinine-blind range”) [10]. However, from data in three GFR segments (above 80, 80 to 40, and below 40 mL/min), they recommended “…observation of subtle changes in serum creatinine levels” at GFRs above 80 mL/min.

We digitized their third figure (mGFR versus sCr) [10], using Graph Grabber v2.0.2 for Windows 10 (Quintessa, Henley-on-Thames, United Kingdom), and plotted the data as solid circles (●), left Y-axis scale in **Fig 8** (see supporting file **S2_dataset**).

**Fig 8.**
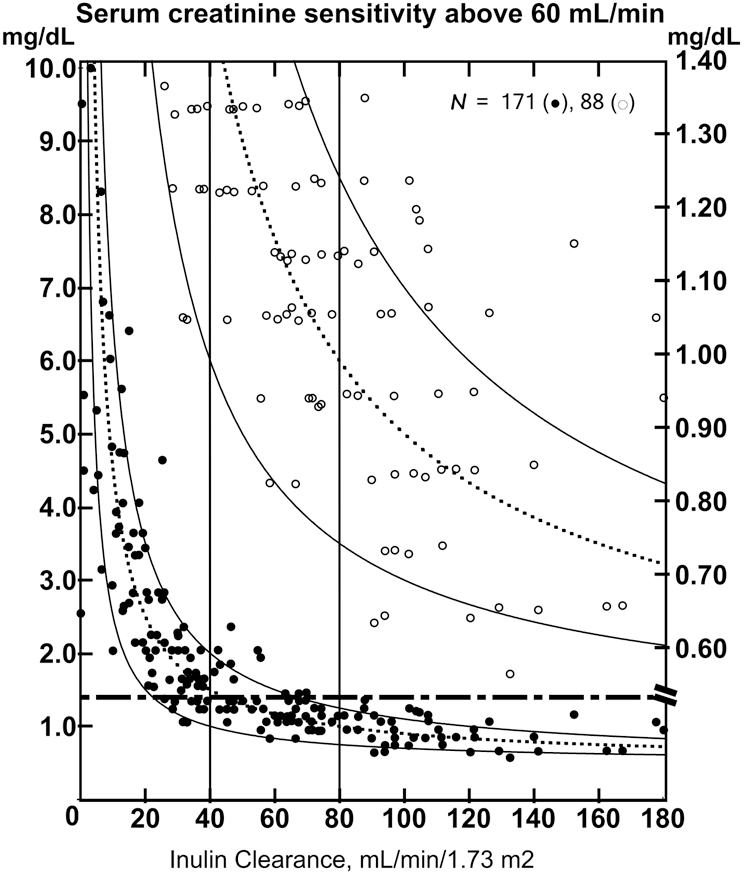
Shemesh et al third figure, sCr versus mGFR. The effect of scale and hyperbola on perception. Solid circles (●) represent 171 patients with glomerular disease, eighty-eight (51%) below the “normal” cutoff—dashed horizontal line at sCr 1.4 mg/dL (124 μmol/L) on the left Y-axis scale. Open circles (○) represent the same patients below the same “normal” cutoff at the top edge on the right Y-axis scale. Overlaying basic hyperbolic curves offered a first visual approximation of the mGFR-sCr relationship. Vertical lines represent boundaries of the three GFR segments.

### 5.2.1 Improving perception with International Units

To better display sCr sensitivity in the preCKD range, we expanded the scale and re-plotted data from the sCr “normal range” as open circles (○), using the right Y-axis scale (downward shifted), **Fig 8**.

Reporting sCr in μmol/L or mg/L units could improve the perception of sCr sensitivity in the preCKD range, but the rough cutoffs alone, in **Fig 1**, may be enough to raise clinician awareness of the implications of small changes in sCr. Modern automated creatinine assays are accurate to 0.05 mg/dL (about 4 μmol/L) or less, and highly precise methods measure differences of 0.03 to 0.04 mg/dL (about 3 to 4 μmol/L) [73].

Enzymatic methods, standardization by isotope-dilution mass spectrometry (IDMS), and other advances further strengthen the recommendation to observe for subtle changes in sCr [10,74]. Lee et al found absolute interlaboratory biases to within ±0.11 mg/dl and relatively constant across the range of sCr, with standard deviations smaller at lower than higher sCr concentrations—0.06 for sCr <1.36 mg/dl (<120 μmol/L, the entire preCKD range) versus 0.10 for sCr >1.36 mg/dl. [75]. Enzymatic assays had less bias than Jaffe assays. There were smaller ranges of difference between highest and lowest sCr value for patients with lower sCr values: the mean ±SD for sCr <1.36 mg/dl (<120 μmol/L) was 0.20 ±0.09 mg/dl, and despite susceptibility to interfering substances, absolute differences between Jaffe and enzymatic results for sCr <1.36 mg/dl (<120 μmol/L) were only 0.05 ±0.07 mg/dL higher. Neubig et al studied paired creatinine values (performed adjacent to each other on the same analytical machine) and found that 75% fell within 5% of equivalence, and 95% fell within 10.3% of equivalence [76].

Hyperbolic functions graphed to zero along the X-axis necessarily compact data along the Y-axis, as Shemesh et al did when plotting all sCr data from 0 to 10 mg/dL (0 to 884 μmol/L) [10]. That compression can suggest little change in sCr along the Y-axis at higher GFRs, above 60 mL/min. However, expanding the Y-axis better matches visual perception to sCr accuracy over the entire preCKD range [77].

The Shemesh et al recommendation that subtle changes in sCr levels can herald changes in GFR [10] was reinforced by subsequent studies correlating small increases in “normal-range” sCr—as little as 0.2 mg/dL (18 μmol/L) compared to the patient’s baseline—with decreased mGFR and more adverse outcomes during long-term follow-up [14,15,21].

However, expanding the scale is helpful but insufficient if the sCr upper reference limit is insensitive. For example, of the 171 glomerulopathic patients in **Fig 8**, 88 (51%) had sCr below the 1.4 mg/dL (124 μmol/L) normal limit, implying Shemesh et al diagnosed them from other signs (e.g., proteinuria) [10]. **Fig 8** shows these patients as solid circles (●) below the horizontal dashed line, left scale, and open circles (○) on the expanded Y-axis, right scale.

### 5.2.2 Modeling Shemesh et al

We hypothesized that modeling Shemesh et al data as a continuous function could extend their observation of using subtle changes in sCr down to 60 mL/min—that the inverse function of kidney filtration and implied sCr and mGFR would better fit their data and allow interpretation with more precision over the preCKD range.

#### Curve fitting

We included all datapoints in the statistical analysis, but to model the relationship of sCr to mGFR, we excluded seven extreme values from fitting the regression curve.

First, we visually fitted a simple hyperbola (dashed line): y = m/x + k, with m = 40, k = 0.5. We derived the regression curve by **(1)** defining a one-sided confidence box (lower limit at ‘slope = minus one’), **(2)** minimizing the sum of squared residuals (squaring the difference between each original data point and corresponding output of best-fit equation), and **(3)** repeating until the cutoff converged to 6.2 mL/min, well below the 80 mL/min dichotomization, for a best-fit curve of m = 39.2, k = 0.60.

Because the inverse relationship between GFR and sCr amplifies errors in the dependent variable when the independent variable is small, this influenced the statistical analysis when deriving sCr (along the Y-axis) from mGFR (along the X-axis), especially as mGFR neared zero, **Fig 8**. Conversely, calculating preCKD eGFRs from the inverse of small values of sCr (e.g., above 60 mL/min) amplifies analytic and physiologic variations of sCr into much larger variance in the eGFR, making eGFR a poor indicator in the preCKD range [78], favoring use of sCr.

#### Residuals

We calculated best-fit residuals (the difference between original datapoint and regression curve), graphed them against mGFR, and assessed their distribution around the X-axis, **Fig 9**. Lack of demographics for individual data points precluded calculating residuals from (or comparing) recent eGFR equations.

**Fig 9.**
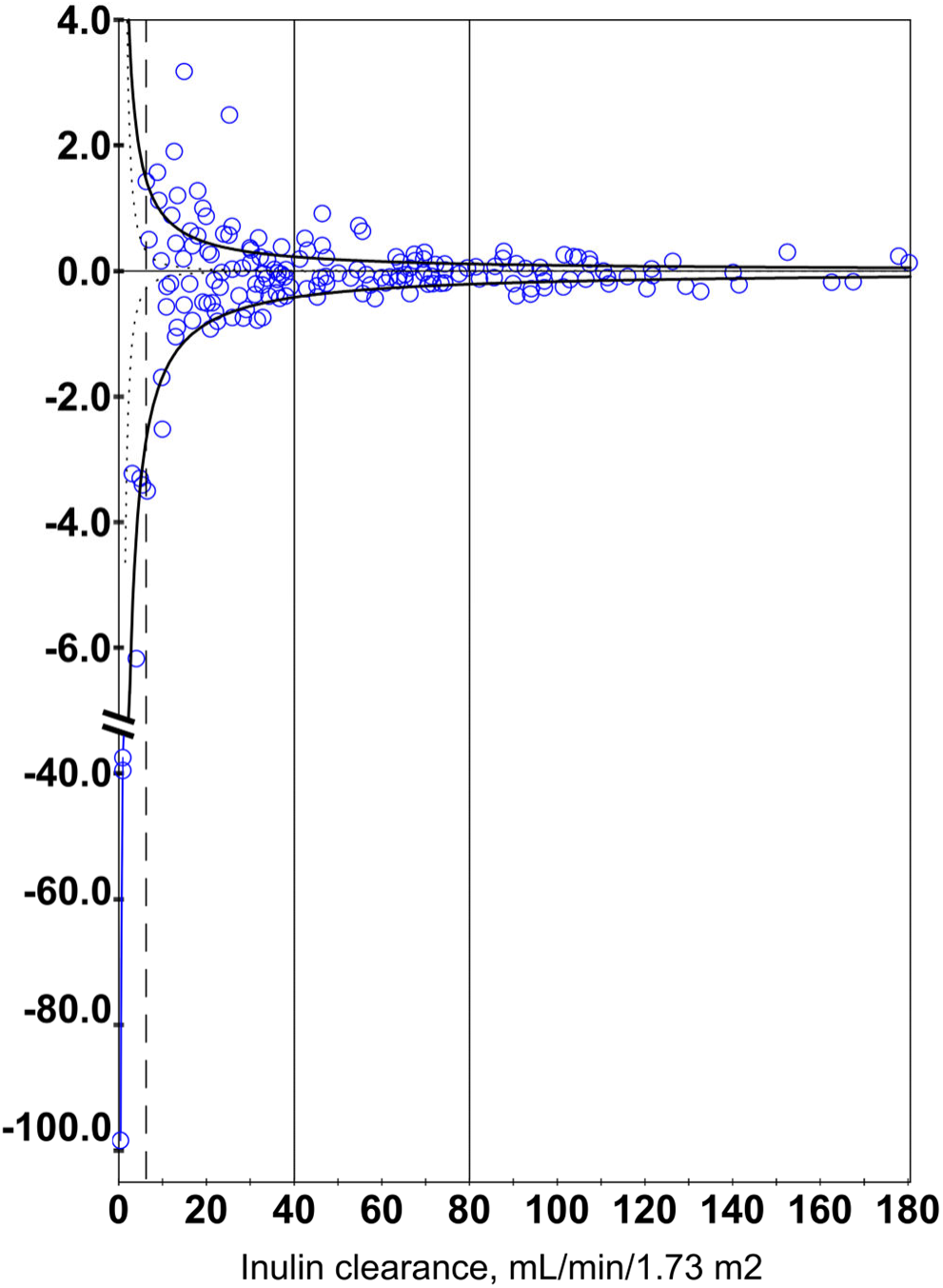
Residuals of Shemesh’s measured sCr minus calculated best-fit sCr. The symmetrical, funnel-shaped variation around the X-axis extends from GFRs of 180 down to the dashed vertical line, indicating the lower limit of the one-sided confidence box—6.2 mL/min—to which the hyperbolic function converged. Dotted black curves represent the limits of sampling error from obtaining data by digitizing figure of Shemesh et al. Large residuals from even small measurement errors at the lower end skewed the statistics (note the Y-axis discontinuity).

Funnel-shaped residuals down to 6.2 mL/min, **Fig 9**, appear equally distributed above and below the X-axis, suggesting random errors, random inter-individual baseline differences, and reasonable agreement between best-fit curve and data over preCKD GFRs. This pattern supports conceptualizing sCr as continuous across the 80 mL/min dichotomized boundary at least down to 60 mL/min, unifying the entire preCKD range.

Nonuniformity (heteroskedasticity) of residuals to the immediate right of 6.2 mL/min may reflect increases in residuals proportional to increases in data values (the hyperbolic function amplifies 30% error bands around each mGFR value, solid black curves).

We used maximum likelihood estimation, which represents the most probable numeric solution for a proposed model, to compare the accuracy of understanding the data either as three segmented groups (sCr below 1.36, between 1.37 and 2.36, or above 2.37 mg/dL—the sample sCr maxima for each GFR segment proposed by Shemesh et al [10]) or as the more conventional continuous inverse function. Although our clinical focus was on GFRs above 60 mL/min, statistical comparison required including the entire mGFR range and all 170 datapoints (one mGFR-sCr pair was not a datapoint because the inverse of zero is mathematically undefined).

Note the marked discontinuity in the Y-axis scale to accommodate three extreme residuals most affected by digitization error at extremely low GFRs. Including these datapoints significantly altered the statistical model equation from our fitted regression curve. Close agreement of fitted and statistical model equations over the preCKD range is not well demonstrated in the model fit statistics, **Fig 10**, that still favor the simpler continuous (inverse/hyperbolic) curve over the dichotomized (grouped) model.

**Fig 10.**
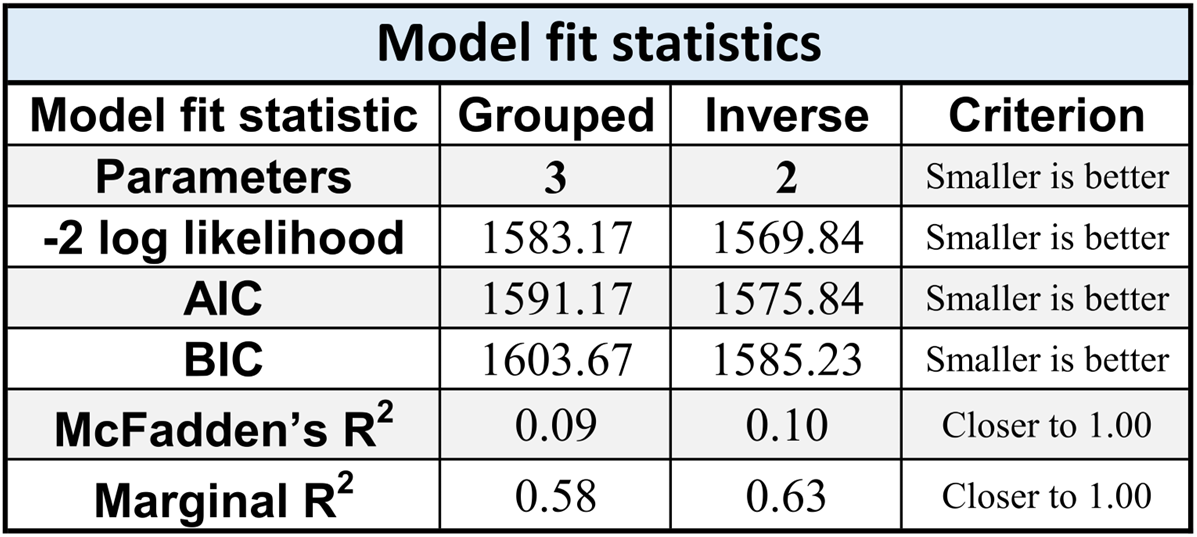
Model fit statistics. The goal of regression modeling is to identify the simplest model that most completely explains the data. The number of parameters represents the number of statistics estimated, with preference given to a model with fewer parameters. The -2 log likelihood estimates the probability of the data given the model, and the AIC and BIC are adjustments to the -2 log likelihood penalizing based on model complexity. A smaller number indicates a better fit of model to data. However, the -2 log likelihood is *X*^2^ distributed and can be used to directly test the comparative fit of two models as the difference of their - 2 log likelihood values with degrees of freedom equal to the difference of their degrees of freedom. Here, the inverse model has significantly better model fit (*X*^2^ (1) = 13.33, *p* = 0.0003). McFadden’s R^2^ and marginal R^2^ both estimate the percentage of variance accounted for by the model from 0.00 (no variance explained) to 1.00 (perfect variance explained). McFadden’s R^2^ is based on the -2 log likelihood relative to a model which only uses the sample mean to explain the data. Marginal R^2^ looks at prediction accuracy, estimated as the correlation between observed GFR scores and predicted GFR scores squared.

Segmenting and dichotomizing continuous measurements [79] introduces ambiguity that confounds dichotomized eGFRs (broken into ranges by arbitrary limits), wherein a slight change in eGFR can mean the difference between CKD and ‘not CKD’.

Our model of continuous function, with each segment equally ‘hyperbolic’, supports incremental comparison to the patient’s baseline. This result is not surprising for a physiologic process but is important to validate use of within-individual change for preCKD. Spanaus et al used a similar approach [14].

### 5.2.3 Tubular secretion of creatinine

Serum creatinine is produced, sequestered, metabolized, and excreted [80]. At steady state, these processes are balanced. Production should be stable on clinical confirmation of stable muscle mass [80]. Sequestration and metabolism may be significant in late CKD, when sCr levels are higher, but are insignificant in steady state or preCKD. Excretion includes renal elimination (glomerular filtration plus tubular secretion) and gastrointestinal elimination (insignificant in preCKD).

TScr equals renal elimination (approximated by CrCl) minus glomerular filtration (approximated by mGFR), but the result is limited by combined analytical errors of CrCl and mGFR. Unlike <u>e</u>GFR inaccuracies that are greatest in preCKD, some <u>m</u>GFR inaccuracies worsen in late CKD as declining GFR markedly lengthens collection times of the exogenous filtration markers [81].

In contrast to sCr sensitivity to GFR changes above 80 mL/min, Shemesh et al advised against using sCr <u>alone</u> to monitor GFR in the middle and lower segments: 80 to 40 and below 40 mL/min. They hypothesized that a slow rise of sCr over the middle segment reflected blunting by an accelerating compensatory rise in TScr [10], **Fig 11, Left Panel,** also shown graphically, **Fig 11, Right Panel**. However, the primary data were quite limited (only three data points), and the difference between CrCl and InCl could also be interpreted as relatively stable, with increasing ratio due to decreasing GFR in the denominator as rising sCr and competing molecules saturate its membrane transporter, limiting tubular secretion in the numerator—a mathematical artifact.

**Fig 11.**
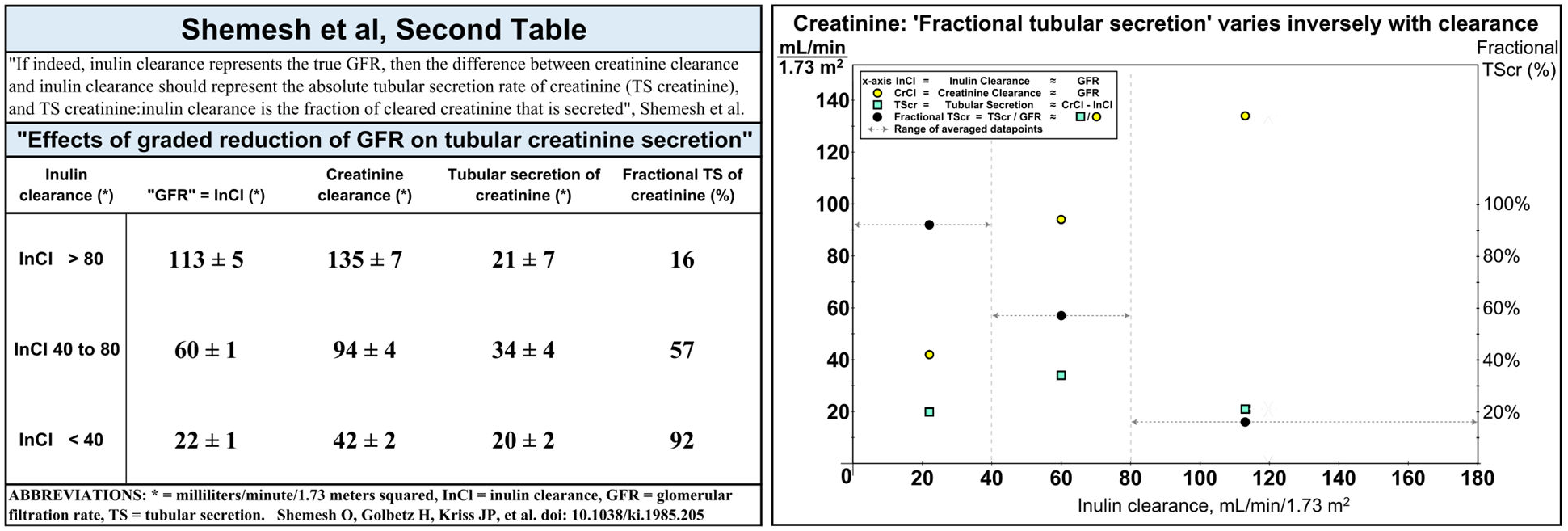
Tubular secretion of creatinine (TScr), after Shemesh et al second table. (left): Because the data is limited (only three data points), TScr could be viewed as relatively stable as GFR, approximated by inulin clearance (InCl), declines. Increasing fractional TScr (the ratio of TScr to InCl) may be largely due to declining InCl in the denominator, **(right):** Displaying the three datapoints graphically suggests the change in TScr over the preCKD range is modest.

We limited our observations to preCKD GFRs to avoid the marked increase, at lower GFRs, in TScr approximated from the difference between CrCl and mGFR <u>when</u> <u>measuring both simultaneousl</u>y [10,82,83]. However, that difference was within the range of measurement error on asynchronous (sequential) measurement (i.e., measuring CrCl before beginning fluid infusion for mGFR) [84], suggesting that intravenous infusions for mGFR may perturb steady-state parameters (perhaps more so in dehydration [40]) that are not affected in isolated measurements of CrCl.

Discoveries since 1985 may improve TScr approximation. Red blood cells sequester enough creatinine in CKD to alter sCr [85,86], with membrane transport that is saturable and susceptible to competitive and noncompetitive inhibition [87]. Cimetidine and other drugs (e.g., trimethoprim, probenecid, cobicistat) inhibit creatinine membrane transporters in renal <u>and</u> extrarenal tissues [88,89,90] (i.e., inhibition of extrarenal sequestration may increase sCr, confounding assumptions about TScr). Further research might show how non-steady-state sCr [91] affects TScr.

More important for prevention, the earliest decreases in GFR occur in the upper segment, above 80 mL/min, where fractional TScr averaged only 16%—within modern P30 standards for kidney tests. Our re-analysis shows TScr does not significantly increase over the preCKD range.

## 5.3 Classic creatinine cofactors

Classic cofactors in GFR estimating equations include age, sex, weight, body surface area, and ‘race’/nationality. Most change slowly (age increases by one every year, body surface area may vary with weight) or almost not at all (sex), and within-individual referencing further marginalizes these weak cofactors. Elsewhere, we examine ‘race’ and nationality for exposure to socially constructed harms [1].

4.3.1 Age

### Adults

Age is an unreliable proxy for CKD. Rule et al scored biopsy specimens of living kidney donors from zero to four for “senile” nephrosclerosis, correlated the results with eGFR, and concluded that age-related decline in GFR in older adults could not be fully explained by nephrosclerosis [92]. About 37% of their kidney donors aged 18 to 29 already had detectable nephrosclerosis, and 10% of those aged 60 to 69 had none, suggesting that kidney injury begins early but is not inevitable.

By deriving eGFR equations from populations diagnosed with CKD or presumed “normal” but silently accumulating kidney injury with age (i.e., unable to detect preCKD), researchers unwittingly normalized age-related GFR decline, as noted by Kallner [93]. Holding sCr steady (e.g., at 1.24 mg/dL or 110 µmol/L) reveals age-related decline in eGFR equations, **Fig 12**. In graphs of longitudinal eGFR versus age, Baba et al found one-quarter of 45,000 “healthy subjects” reached the cutoff for CKD stage G3 by age 75 but maintained relative chart positions because all showed age-related decline [94]. In retrospective examination of 47,000 hospital admissions, Xu et al showed the risk of AKI increased only *after* age 75 years [95]. The eGFR yields high rates of senile “CKD” in otherwise healthy elderly [78,96,97,98]—an estimated over-diagnosis of CKD in the elderly by 30 to 50% [99].

**Fig 12.**
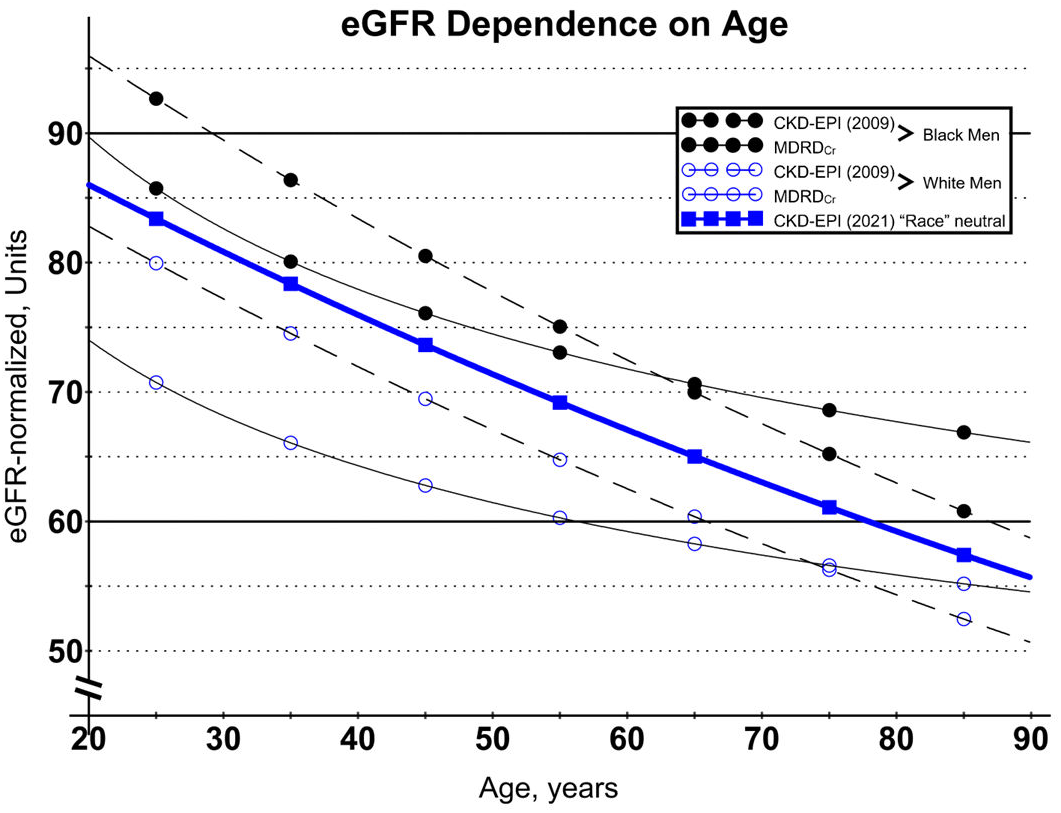
Stable creatinine reveals decline in GFR versus age and the effect of ‘race’ in eGFR equations. Holding sCr steady at 110 μmol/L (1.24 mg/dL, the boundary between pre-CKD and CKD stage G3) demonstrates eGFR dependence on age. The graph shows MDRD_Cr_, CKD-EPI (2009), and CKD-EPI (2021) “race neutral” eGFR equations versus age for Black and White men. The horizontal lines at 90 and 60 mL/min represent the boundaries between CKD stages 1 and 2 and CKD stages 2 and 3. The graph may explain overdiagnosis of CKD in the elderly (see text and discussion at Kallner’s second figure [52]). When ‘race’ is misused in pre-test “population weighting,” the CKD-EPI (2021) “race neutral” equation can alter CKD diagnosis through post-test shifting of results from Black and White patients, respectively, above and below the thick blue line (see text). Note: clinical use often omits normalization of eGFR to body surface area.

Self-referenced longitudinal sCr could redirect resources from healthy elderly to prevention services for younger patients with pre-CKD. Tempering eGFR results, Morgan et al observed:

> … to assess a patient’s glomerular function, one does not really need to know the GFR but rather whether the GFR is appropriate for the patient’s age and physique, and thus whether he is able to maintain his internal environment within acceptable limits…. a normal P [plasma creatinine] can be taken as evidence that a patient’s glomerular function is adequate for the demand on it, whether or not it has changed as a result of disease [100].

### Pediatrics

The Schwartz equation for pediatric eGFR estimation was similarly derived from children with CKD (stages 2 to 4), so it is unlikely to separate normal from pre-CKD. Gretz suggested tracking pediatric CKD with “progression charts” of direct markers (e.g., sCr) versus time [11]. If the developmental sCr chart can reveal preCKD, it may allow research to identify and prevent the earliest kidney injuries.

However, dynamic changes from birth to 20 years complicate interpretation of pediatric sCr charts, which can be roughly divided into three epochs—0 to 3 weeks, 3 weeks to 3 years, and 3 to 20 years—to accommodate three developmental processes that affect sCr:

**(1)** loss of fetal-maternal-placental equilibration with clearance by maternal kidneys (first 3 weeks),
**(2)** kidney growth increases GFR from ∼20 mL/min at birth to ∼120 mL/min (first 3 years), and
**(3)** muscle growth increases endogenous creatinine production (first 20 years) [101].

In the first 3 weeks, kidney and muscle growth are negligible, allowing sCr modeling to emphasize the decay in fetal-maternal-placental equilibration. In the first 3 years, sCr modeling requires approximation of kidney size (e.g., kidney length) and muscle mass (e.g., patient height). From 3 to 20 years, enlarging muscle mass is the predominant developmental factor.

### 5.3.2 Sex hormones

Adjustments for women typically reference population differences in size and muscularity, likely incorporating conscious and unconscious stereotypes of occupation and lifestyle. However, individual women are increasingly athletic (especially since Title IX) and defy a uniform ‘correction’ for physical activity [37].

Blood levels or specific medical circumstances (e.g., oophorectomy, hormone replacement therapy, gender reassignment) may capture the effects of sex hormones. Reports suggest potential positive (decreased kidney fibrosis) and negative (increased CKD) effects of estrogen and estrogen receptors in the kidney [102,103,104].

### 5.3.3 Body surface area, BMI

Indexing kidney measurements to body surface area had little effect on normal body size populations but introduced significant bias at extremes of weight—obese and anorectic patients [105,106]. Examining increased AKI risk during COVID using the Edmonton Obesity Staging System (EOSS) showed “…elevated risk is driven by obesity-related comorbidity burden, not BMI” [107]. Begley noted, “Obesity is a marker of poverty…. a signal for so many of the social determinants of health” [108]. These reports favor absolute, non-indexed kidney measurements.

## ACKNOWLEDGMENTS

In memory of Rear Admiral W. Norman Johnson and many others who endured kidney failure after prescribed nephrotoxic drugs that might have been avoided with early warning and caution, and of the Rev. Dr. Canon Cyril C. Burke, Sr, who taught ethics and whose final medical care was complicated by ‘race’.

The authors gratefully acknowledge Edward Feller, MD, FACP, FACG, Ruth Levy Guyer, PhD, and anonymous colleagues for critical review of a draft of this article; and Joseph J. Fins, MD, MACP, FRCP, and John C. Kotelly, PhD, US Air Force mathematician (ret.), for their insights.

## ABBREVIATIONS

ASC: adult serum creatinine
CKD: chronic kidney disease
CrCl: creatinine clearance
eGFR: estimated GFR
GFR: glomerular filtration rate
KF: kidney failure
m^2^: square meters
mGFR: measured GFR
mg/dL: milligrams per deciliter
mL/min: milliliter per minute
μmol/L: micromoles per liter
NSAID: nonsteroidal anti-inflammatory drugs
P30: percentage of estimates within ±30%
RCV: reference change value
sCr: serum creatinine
sCrMax: maximum sCr to date
sCrRCV: serum creatinine reference change value
sCysC: serum cystatin C
TScr: tubular secretion of creatinine.

